# Antibiotic Use and Outcomes in Young Children Hospitalized with Uncomplicated Community-Acquired Pneumonia

**DOI:** 10.1101/2021.06.18.21259114

**Authors:** Meghan E. Hofto, Nichole Samuy, Robert F. Pass

## Abstract

**Objectives:** To compare children aged 36 months or younger hospitalized with uncomplicated community-acquired pneumonia (CAP) that are not treated with antibiotics to those treated with antibiotics in terms of clinical features and outcome measures.

**Methods:** Administrative data and medical record review were used to identify patients from 3-36 months of age hospitalized from 2011-2019 with uncomplicated CAP. Patients were considered treated if they received antibiotics for more than 2 inpatient days and/or at discharge, and not treated if they received 2 or fewer inpatient days and no antibiotics at discharge. Demographic features, clinical characteristics, radiologic findings, viral testing, illness severity, length of stay and 30-day hospital readmissions were assessed and compared according to antibiotic treatment.

**Results:** 322 CAP cases were included. 266/322 (83%) received antibiotics for more than 48 hours and/or at discharge. 56 patients received 2 or fewer inpatient days of antibiotics and no antibiotics at discharge; the majority received no inpatient antibiotics. There were no differences between the two groups in illness severity, length of stay or hospital readmissions. The proportion of patients treated with antibiotics decreased from 88% (2011-2013) to 66% during the most recent years studied (2017-2019).

**Conclusion:** There was no difference in outcome of uncomplicated CAP in previously healthy children less than 36 months of age between those treated and not treated with antibiotics. Additional tools are needed to facilitate identification of viral CAP in young children and decrease unnecessary antibiotic use.

## INTRODUCTION

Although it is generally accepted that viruses cause the majority of community-acquired pneumonia (CAP) in young children, narrow spectrum antibiotics (typically penicillin, amoxicillin, or amoxicillin/clavulanate) are recommended for all children hospitalized with uncomplicated CAP by multiple guidelines promulgated by subspecialty societies and health service systems.^1-5^ While monitoring adherence to guideline recommendations for use of narrow spectrum antibiotics for children hospitalized with CAP, we noted that some patients did not receive antibiotic treatment. The purpose of this study was to determine the proportion of children aged 3 months to 3 years hospitalized with CAP that did not receive a treatment course of antibiotics and assess any differences in clinical outcome related to treatment.

## METHODS

### Study Design

This was a retrospective observational study of children with uncomplicated CAP hospitalized between September 2011 and December 2019 at a single, freestanding children’s hospital with 380 beds and over 15,000 admissions per year. The primary objective of this study was to determine the proportion of hospitalized children aged 3 to 36 months treated with antibiotics for uncomplicated CAP. The secondary objective was to determine if patients not treated with antibiotics worsened during hospitalization, requiring intensive care, or more frequently required readmission after discharge. This study used the definition of CAP (“signs and symptoms of pneumonia in a previously healthy child caused by an infection that has been acquired outside of the hospital”) included in the Infectious Diseases Society of America/Pediatric Infectious Diseases Society guideline.^3^ The Institutional Review Board of the University of Alabama at Birmingham approved this study.

### Study Population

Children admitted with an *International Classification of Disease, Ninth Revision* (ICD-9) or an *International Classification of Disease, Tenth Revision* (ICD-10) discharge diagnosis code for pneumonia (480-487.0 or J12.0-J18.9, respectively) were identified for possible inclusion. The target population was children 3 to 36 months of age without underlying comorbidities that increase the risk of bacterial pneumonia or cause acute respiratory symptoms independent of infection. Patients with chronic lung disease (other than asthma), immunodeficiency, admission for status asthmaticus, or prior admission within one month of the index hospitalization were excluded. Patients with a clinical diagnosis of aspiration pneumonia or prior history of aspiration in the medical record were also excluded. Patients were excluded if they had complicated CAP (“parapneumonic effusions, multilobar disease, abscesses or cavities, necrotizing pneumonia, empyema, pneumothorax, or bronchopleural fistula” per the guideline^3^) or received treatment with antibiotics for a concomitant infection other than CAP.

Administrative data, including age, sex, race/ethnicity, length of stay and discharge diagnosis codes, were obtained from the local institution. Diagnosis codes were first reviewed, then chart review on identified patients was performed to confirm that they met the guideline definition of CAP. Microbiologic results, including blood cultures and viral polymerase chain reaction (PCR) testing, 30-day readmissions, as well as antimicrobial use before, during and after hospitalization were obtained from medical record review. Radiographic results were evaluated based on the radiologist read of the image and categorized as “focal opacity” if “pneumonia,” “cannot exclude pneumonia” or “focal opacity” were mentioned, or “viral versus atelectasis” if there was a mention of presumed viral process or atelectasis. There was overlap between the two categories, as some reports mentioned “likely viral process or atelectasis but cannot rule out developing pneumonia”. The “treated with antibiotics group” (TAG) was defined as the patients treated inpatient for more than 2 days (48 hours) with antibiotics or discharged home with a prescription for an antibiotic, and the “no antibiotics group” (NAG) was defined as the group treated inpatient for 2 days or less and discharged home on no antibiotics. These groups were defined prior to initiation of this study. We selected these specifications because we thought they would accurately reflect a clinical decision to treat for possible bacterial infection (TAG) or not to treat (NAG). The possibility of overlap between the two groups in antibiotic use was recognized.

### Statistical Analysis

Statistical analysis was performed using commercially available software (SAS 9.4; SAS Institute Inc, Cary, NC). Proportions were compared using Fisher’s Exact test and Chi Square. Continuous variables were analyzed using Wilcoxon rank sum test. P values < 0.05 were considered significant.

## RESULTS

### Identification of the Study Population

Administrative data identified 2,313 patients aged 3 months to 19 years with possible CAP based on a discharge diagnosis code. Review of diagnosis and procedure codes excluded 997 (43%), leaving 1,316 cases. Medical record review excluded an additional 338. Figure 1 details the reasons for exclusion. Of the remaining 978 patients, 444 patients were aged 3 months to 36 months; 15 were excluded because they received antibiotics for another indication (14 for acute otitis media, 1 for urinary tract infection), and 107 were excluded due to complicated CAP. All CAP complications were either present on admission or developed in patients already on antibiotics.

**Figure 1.**
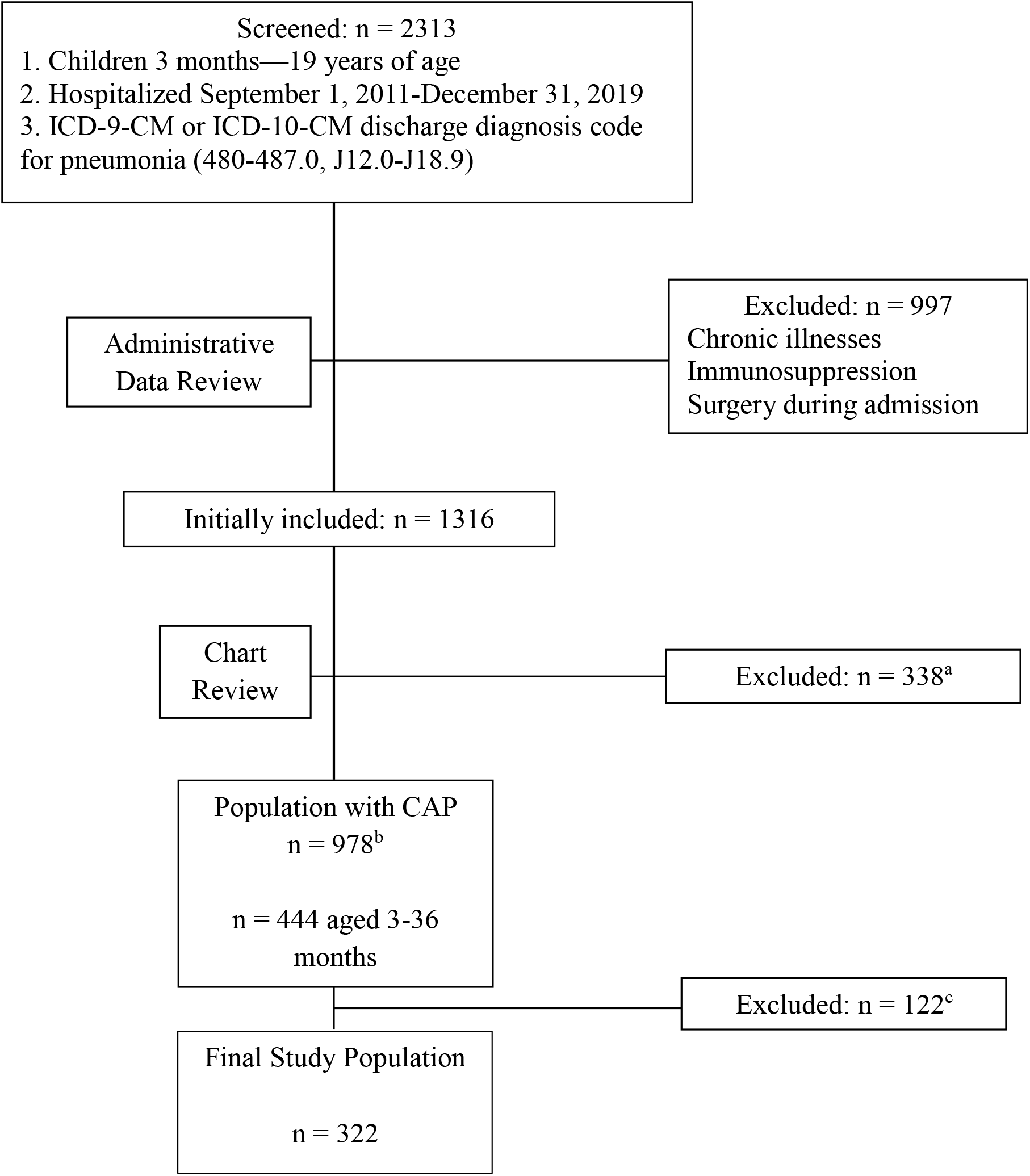
Accrual of the study population.

### Demographic and Clinical Characteristics of the Study Population

Of the 322 patients with uncomplicated CAP in the final study population, 266 (83%) were treated with or discharged home on antibiotics. Table 1 shows demographic and clinical characteristics comparing the TAG to the NAG. The NAG was older (25 months versus 17 months, P = 0.003) than the TAG. There were no statistically significant differences in sex, race, or ethnicity between the groups. Fever, hypoxemia at time of admission, and vomiting were present at similar rates, with a similar maximum pre-admission temperature recorded. The TAG was more likely to have received antibiotics outpatient or in the Emergency Department (236/266, 89%) and on admission (263/266, 99%) than the NAG (37/56, 66%, prior and 24/56, 43%, after admission), P < 0.001 for both comparisons.

**Table 1.** Characteristics of patients with community-acquired pneumonia (CAP) categorized by presence or absence of antibiotic treatment.

|  | **Treated with**<br>**Antibiotics Group**<br>**N = 266** | **No Antibiotic Group**<br>**N=56** | **P value** |
| --- | --- | --- | --- |
| Sex, female | 124 (47)<sup>a</sup> | 20 (36) | 0.1 |
| Race/Ethnicity, |  |  |  |
| African American | 112 (42) | 16 (29) | 0.07 |
| White Non-Hispanic | 119 (45) | 32 (57) | 0.1 |
| White Hispanic | 31 (12) | 8 (14) | 0.7 |
| Other | 4 (1) | 0 |  |
| Age, median months [IQR] | 17 [11-26] | 25 [16-31] | 0.003 |
| Fever prior to admission | 232 (87) | 45 (80) | 0.2 |
| Tmax<sup>b</sup> (°F), median [IQR] | 103 [102-104] | 103 [101.2-103.6] | 0.4 |
| Hypoxemia | 108 (41) | 24 (43) | 0.8 |
| Vomiting | 130 (49) | 25 (45) | 0.6 |
| Antibiotic Use |  |  |  |
| Prior to admission<sup>c</sup> | 236 (89) | 37 (66) | < 0.001 |
| After admission | 263 (99)<sup>d</sup> | 24 (43) | < 0.001 |
| Intensive Care Unit | 11 (4) | 2 (4) | 1 |
| Length of stay, median days [IQR] | 2 [1-3] | 1.5 [1-3] | 0.3 |
| Readmissions<sup>e</sup> | 13 (5) | 2 (4) | 1 |

<sup>a</sup>Numbers in parentheses are percent.
<sup>b</sup>Maximum temperature if febrile prior to admission by parental report or Emergency Department record.
<sup>c</sup>Includes antibiotics prescribed prior to the Emergency Department and those given in the Emergency Department.
<sup>d</sup>3 patients did not receive antibiotics after admission, but they all stayed less than 24 hours and were prescribed antibiotics at discharge.
<sup>e</sup>30 day readmissions to the inpatient setting.

Outcome measures were similar between the two groups, Table 1. Similar proportions of patients in both groups required care in the intensive care unit (ICU) during hospitalization, and all but 2 patients (both in the TAG) requiring the ICU were initially admitted to the ICU. There were no statistically significant differences between the TAG and the NAG in either hospital length of stay or 30 day readmissions. Both readmissions in the NAG were due to new viral illnesses after complete recovery from the index hospitalization. Of the 13 TAG readmissions, seven were due to persistent or worsening symptoms, and six were due to new illness after complete recovery from the index hospitalization.

### Radiologic and Laboratory Findings

The majority (311/322, 97%) of patients had radiographic imaging during hospitalization. The NAG was more likely to have a radiologist read of viral process or atelectasis (50/51, 98%) than the TAG (178/260, 68%), P < 0.001. The TAG was more likely to have a read suggesting possible bacterial pneumonia than the NAG, (182/260, 70%, versus 8/51, 16%, P < 0.001). There was considerable overlap in radiologist impressions with mention of both viral airways disease and pneumonia or focal opacity, more frequently observed in the TAG, 100/260 (38%) than in the NAG, 7/51 (14%), P = 0.01.

The median (interquartile range) white blood cell count (x 10^9^/L) was 12.0 (8.7 – 19.0) for the TAG and 10.7 (7.8 – 14.1) for the NAG, a difference that was not statistically significant. Fifty-five percent of the TAG and 58% of the NAG had blood cultures obtained. All blood cultures were negative.

Viral testing was more common in the NAG, 39/56 (70%) versus the TAG, 152/266 (57%), P = 0.01, Table 2. Forty-eight percent (73/152) of the TAG was positive for at least one virus compared with 67% (26/39) of the NAG (P = 0.001). For both groups, respiratory syncytial viruses (RSV) were the most commonly detected, followed by influenza viruses. Seven patients (5 in the TAG and 2 in the NAG) had two viruses detected on testing. Of the 73 TAG patients positive for a virus, there were no significant differences from other TAG patients in maximum pre-admission temperature, age, chest radiograph findings, or hypoxemia; these patients were more likely to be treated in the ICU than other TAG patients (8/73, 11%, vs 3/193, 2%, p = 0.002).

**Table 2.** Virus detection, including multiplex viral polymerase chain reaction, rapid influenza, and rapid respiratory syncytial virus (RSV) testing by group.

|  | **Treated with**<br>**Antibiotics Group**<br>**N = 266** | **No Antibiotic Group**<br>**N=56** | **P value** |
| --- | --- | --- | --- |
| Positive | 73 (27)<sup>a</sup> | 26 (47) | 0.001 |
| Negative | 79 (30) | 13 (23) |  |
| Not obtained | 114 (43) | 17 (30) |  |
| Respiratory Syncytial<br>Virus | 34 (44)<sup>b</sup> | 13 (45)<sup>c</sup> | 1 |
| Influenza | 15 (19) | 6 (21) | 1 |
| Human Metapneumovirus | 11 (14) | 3 (10) | 0.8 |
| Rhinovirus/Enterovirus | 7 (9)<sup>d</sup> | 2 (7)<sup>e</sup> | 1 |
| Parainfluenza | 5 (6) | 3 (10) | 0.7 |
| Adenovirus | 6 (6) | 2 (7) | 1 |

<sup>a</sup>Numbers in parentheses are percent.
<sup>b</sup>Including RSV + adenovirus (1) and rhinovirus/enterovirus (2)
<sup>c</sup>Including RSV + influenza (1)
<sup>d</sup>Including rhinovirus/enterovirus + human metapneumovirus (1), parainfluenza (1), RSV (2)
<sup>e</sup>Including rhinovirus/enterovirus + adenovirus (1)

### Antibiotic Exposure in the TAG and the NAG

We examined the data for possible overlap in antibiotic exposure between the TAG and NAG. Among the TAG, 257/266 (97%) were discharged from the hospital with an antibiotic prescription. Seven of the 9 not discharged on antibiotics completed 6 or more days of treatment prior to discharge, and the other two patients had 3 or 4 inpatient days of treatment plus prehospital treatment. In contrast, no NAG patients were prescribed antibiotics at discharge; 32 of them received no inpatient antibiotics. Among the 24 NAG patients that received ≤ 2 days of inpatient antibiotics, 11 received ≤ 1 day of inpatient antibiotics and ≤ 1 dose of antibiotics prior to hospitalization. Five NAG patients received a single dose of antibiotics prior to admission and had antibiotics discontinued on hospital day 2. Among the remaining 8 NAG patients, there were only 2 that received 2 inpatient days of antibiotics and had more than 4 total days of antibiotics prior to hospitalization. We cannot exclude the possibility that treating physicians concluded that those 2 NAG patients were thought to be fully treated, but both were diagnosed with viral pneumonia in the medical record when antibiotics were discontinued. Narrow spectrum antibiotics were used to treat 175 (66%) of TAG patients and 50% of the 24 NAG patients that received antibiotics.

### Trends in Antibiotic Use

There were no significant differences in antibiotic use at discharge by quarter of the year; 85% of those discharged in quarters 2 and 3 (local Spring and Summer) and 81% discharged in quarters 1 and 4 (local respiratory virus season) were prescribed antibiotics at discharge. Antibiotic use significantly declined in recent years. The TAG contained 88% (134/152) of patients from 2011-2013, 86% (83/96) of patients from 2014-2016, and only 66% (49/74) of patients from 2017-2019 (P < 0.001).

## DISCUSSION

In this study of 322 healthy children aged 36 months or less hospitalized with uncomplicated CAP, there were 56 that did not receive a treatment course of antibiotics. While there were differences in the radiographic findings and age, there were no other differences between those patients discharged on antibiotics and those that were not, in either indicators of disease severity (fever, hypoxemia, vomiting, ICU stay) or in other demographic features. There was no difference between TAG and NAG patients in length of hospital stay or hospital readmissions. No patient from the NAG required readmission for antibiotic treatment. In addition, the proportion of patients treated with antibiotics decreased from 88% (2011-2013) to 66% during the most recent years studied (2017-2019), evidence that it is possible to identify patients less than 3 years of age hospitalized with uncomplicated CAP that do not require antibiotic treatment.

There have been multiple studies in recent years providing evidence that CAP in this age group is predominantly caused by viruses.^6-10^ A 2010 study reported rates of PCR detection of respiratory viruses in 90% of children less than 3 years of age hospitalized with pneumonia and 52% in a control group.^10^ A study conducted in hospitals in 3 U.S. cities using molecular methods reported detection of respiratory viruses and bacteria respectively in 66% and 17% of 2222 children with CAP; the preponderance of viruses was more striking in younger children.^7^ Detection of respiratory viruses other than rhinovirus occurred in less than 3% of a control group. A 2017 paper from the same research group focused on viruses and reported respiratory virus detection in 69% of children hospitalized with CAP and 24% of asymptomatic controls.^9^ A study from Sweden that compared viral detection rates in children with CAP and controls reported significantly higher rates of infection with influenza, human metapneumovirus and RSV in cases.^8^ Similar results were reported in a study from Australia with strikingly increased rates of infection in cases compared with controls for RSV, human metapneumovirus, influenza viruses and adenovirus.^6^ Based on these and other reports, the evidence that the vast majority of uncomplicated CAP cases in hospitalized young children are due to viruses, not bacteria, is persuasive.

We found that physicians were increasingly willing to overlook guideline recommendations for treating all hospitalized children with CAP with antibiotics. It is possible that growing awareness of studies using molecular methods to determine pneumonia etiology influenced physician management. There were no active efforts at the study hospital to encourage decreased use of antibiotics for hospitalized children with uncomplicated CAP. Comparing NAG to TAG patients provided some suggestions as to data affecting clinical decision-making. Focal finding on chest radiograph and antibiotics started on admission were more common in the TAG. Mention of viral process or atelectasis and a positive result on viral testing were associated with a diagnosis of viral pneumonia and no antibiotic treatment. There were no statistically significant differences in antibiotic use by detected virus.

Chest radiographs were obtained in almost every patient in the current study, and misinterpretation of atelectasis as a focus of bacterial infection on chest radiograph could be driving antibiotic use. The American Academy of Pediatrics Guideline on Bronchiolitis recommends avoiding chest radiographs.^11^ Given the overlap between pneumonia and bronchiolitis in younger children, decreasing imaging in bronchiolitis could decrease unnecessary antibiotic use.

Studies of CAP in children and adults report that measuring serum levels of C-reactive protein and/or procalcitonin may assist in distinguishing bacterial from viral respiratory infections.^12,13^ Wider use of these tests in young children hospitalized with pneumonia may possibly lead to a decrease in unnecessary use of antibiotics. However, neither of these tests were used consistently in the study population, and routine use of these tests has not been endorsed by guidelines. Improved means of identifying children with CAP that do not require antibiotic treatment will have a significant impact on antibiotic stewardship and preventing harm, as antibiotic-associated adverse events occur frequently in hospitalized children and require additional medical interventions.^14^

It is important to recognize that the current study was limited to previously healthy inpatients with uncomplicated CAP between 3 and 36 months of age, without recent hospitalizations, chronic lung disease or immune impairment due to disease or medications. Results should not be used to inform treatment of pneumonia patients that do not meet those criteria. A strength of this study is that we reviewed medical records to confirm that included patients met the guideline definition of CAP and to collect clinical data. We acknowledge limitations of this study. It was retrospective; a prospective, protocol driven study would be able to collect more clinical and laboratory findings and study physician decision making. This study was conducted at a single site and is unable to assess hospital based or regional differences in antibiotic prescribing practices for CAP.

## CONCLUSIONS

Most pneumonia in children less than three years of age is due to viral infection, and recent studies have challenged the idea that antibiotics are necessary in younger patients with CAP.^15-17^ Based on the current study, it appears that a substantial proportion of previously healthy children less than 3 years of age hospitalized with uncomplicated CAP will do well without antibiotic treatment. Better tools for identifying those that require antibiotics will make it possible to achieve a significant advance in antibiotic stewardship.

## Data Availability

Data available on request.

## Abbreviations

(CAP): community-acquired pneumonia
(ICD-9): *International Classification of Disease, Ninth Revision*
(ICD-10): *International Classification of Disease, Tenth Revision*
(TAG): “treated with antibiotics group”
(NAG): “no antibiotics group”
(ICU): intensive care unit
(RSV): respiratory syncytial virus
(PCR): polymerase chain reaction

